# Unemployment insurance and food insecurity among people who lost employment in the wake of COVID-19

**DOI:** 10.1101/2020.07.28.20163618

**Authors:** Julia Raifman, Jacob Bor, Atheendar Venkataramani

## Abstract

Forty million U.S. residents lost their jobs in the first two months of the coronavirus disease 2019 (COVID-19) pandemic. In response, the Federal Government expanded unemployment insurance benefits in both size ($600/week supplement) and scope (to include caregivers and self-employed workers). We assessed the relationship between unemployment insurance and food insecurity among people who lost their jobs during the COVID-19 pandemic in the period when the federal unemployment insurance supplement was in place. We analyzed data from the Understanding Coronavirus in America (UAC) cohort, a longitudinal survey collected by the University of Southern California Center for Economic and Social Research (CESR) every two weeks between April 1 and July 8, 2020. We limited the sample to individuals living in households earning less than $75,000 in February 2020 who lost their jobs during COVID-19. Using difference-in-differences and event study regression models, we evaluated the association between receipt of unemployment insurance and self-reported food insecurity and eating less due to financial constraints. We found that 40.5% of those living in households earning less than $75,000 and employed in February 2020 experienced unemployment during the COVID-19 pandemic. Of those who lost their jobs, 31% reported food insecurity and 33% reported eating less due to financial constraints. Food insecurity peaked in April 2020 and declined over time, but began to increase again among people receiving unemployment insurance during the final wave of the survey ahead of the federal supplement to unemployment insurance ending. Food insecurity and eating less were more common among people who were non-White, lived in lower-income households, younger, and who were sexual or gender minorities. Receipt of unemployment insurance was associated with a 4.4 percentage point (95% CI: −7.8 to −0.9 percentage points) decline in food insecurity (a 30.3% relative decline compared to the average level of food insecurity during the study period). Receipt of unemployment insurance was also associated with a 6.1 percentage point (95% CI: −9.6 to −2.7 percentage point) decline in eating less due to financial constraints (a 42% relative decline). Estimates from event study specifications revealed that reductions in food insecurity and eating less were greatest in the four-week period immediately following receipt of unemployment insurance, with no evidence of differential pre-existing trends in either outcome. We conclude that receiving unemployment insurance benefits during the period when the $600/week federal supplement was in place was associated with large reductions in food insecurity.

## Introduction

The impacts of Coronavirus Disease 2019 (COVID-19) extend well beyond morbidity and mortality. In the United States, COVID-19-related business closures and reductions in economic activity have led to a sharp rise in unemployment rates, from 3.5% in February 2020 to 14.7% in April 2020. As of June 2020, the unemployment rate stands at 11.1%.^1^ Job losses over this period have been concentrated among people living in low-income households, and resulting drops in income have made many individuals and families vulnerable to food insecurity.^2,3^ Food insecurity is defined by the U.S. Department of Agriculture as “household-level economic and social condition of limited or uncertain access to adequate food.” Food insecurity is associated with worse general health and well-being,^4–6^ physical hunger pangs and fatigue, psychological depression, anxiety, suicidal ideation,^7^ and interpersonal stress and challenges,^6,8–10^ as well as chronic disease,^5^ and worse developmental outcomes for children.^4,11^ Initial evidence suggests that food insecurity has more than doubled among all households and tripled among households with children during the COVID-19 pandemic relative to February 2020.^12^

Unemployment insurance has taken on critical importance in the wake of the rapid rise in unemployment and in expectation of a COVID-19-related recession. The federal Coronavirus Aid, Relief, and Economic Security Act (CARES Act) authorized a $600/week federal supplement to weekly state unemployment benefits until July 2020. In 48 states, the $600 federal supplement has brought unemployment insurance benefits up to or above a living wage for a single adult or a family of two working adults with two children^13^ (Massachusetts and Washington unemployment insurance amounts provide a living wage). The CARES Act also created the Pandemic Unemployment Assistance (PUA) program to cover independent contractors, people advised by their health care providers to quarantine due to pre-existing health conditions that may raise the risk of critical or death illness from COVID-19, and people unable to work due to long-term health consequences of COVID-19 or caregiving responsibilities related to COVID-19. The Pandemic Emergency Unemployment Compensation (PEUC) program extended the duration of unemployment benefits coverage for 13 additional weeks after state unemployment insurance payments end. In addition to federal legislation, many states also acted to expand unemployment eligibility or benefits or to relax requirements. By July 2020, 44 states and the District of Columbia (DC) had expanded unemployment insurance eligibility or duration in some capacity, and 36 of the 41 states with work search requirements for unemployment insurance eliminated the requirement.^14^ Prior evidence indicates that unemployment insurance was associated with reduced food insecurity^15^ and reduced suicide rates.^16^

With the $600/week federal supplement to unemployment insurance set to expire, there is growing concern that low-income households will face even greater financial and food insecurity in the coming months, particularly as COVID-19 case rates have increased and unemployment rates remain at historically high levels. At this writing, Congress is considering whether to extend the supplement to unemployment insurance. States will also continue to consider whether to expand unemployment insurance benefits. These decisions will have important consequences for food insecurity. To help inform policy decisions, we evaluated the relationship between unemployment insurance and food insecurity among people who lost their jobs between April to July 2020, a period during which unemployment benefits were elevated relative to historic levels. We hypothesized that unemployment insurance would have a particularly large effect on food insecurity because, with the $600 CARES Act supplement, it provided routine financial support in a sufficient amount to support basic needs such as food and housing.

## Methods

### Data

We used data from the University of Southern California’s Center for Economic and Social Research (CESR)’s Understanding Coronavirus in America (UCA) study.^17^ The UCA is an extension of an ongoing Internet-based, longitudinal research survey with approximately 8500 participants.^18^ Beginning on April 1, 2020, CESR began conducting repeated UCA surveys on health and economic outcomes with the full panel every two weeks (Appendix Table 1). CESR randomly assigned panel members to be invited to respond on a pre-assigned day of a two-week period, so that the full sample was invited to participate over each 14-day period. Participants received an extra monetary incentive to complete the survey on the day they were invited to participate. The data are made publicly available to inform rapid response research and reporting. We used data collected over 6 waves, between April 1 and July 8, 2020.

### Sample

We restricted the sample to people living in households earning less than $75,000 in the past 12 months (to focus on the population most at risk for food insecurity); who participated in at least 2 survey waves; who reported being employed in February 2020, prior to the COVID-19 pandemic; and who lost employment at some point during the April to July study period. We adopted the last sample restriction because we anticipated that trends in food insecurity would differ among those who remained employed.

We determined household income based on data collected at baseline. Survey participants were asked, “Which category represents the total combined income of all members of your family (living in your house) during the past 12 months? This includes money from jobs, net income from business, farm or rent, pensions, dividends, interest, Social Security payments and any other monetary income received by members of your family who are 15 years of age or older.” We considered individuals to have been employed in February based on their answers to the question, “Thinking back to February 2020, were you employed by the government, employed by a private company, employed by a nonprofit organization, self-employed, not employed or retired?” We considered people to have been employed in February based on any response besides “not employed” or “retired.” Participants who lost their jobs during the study period were identified as those who responded “no” to the question, “Do you currently have a job?” We included in the sample anyone who was employed in February 2020 and who reported being unemployed at any point during the survey.

### Exposure

The primary exposure of interest was beginning to receive unemployment insurance, a binary variable coded as 1 beginning in the first wave in which the respondent answered “yes” to the question “Have you received unemployment insurance benefits in the past fourteen days?” and as 1 thereafter (there were 7 observations excluded because participants reported they were “unsure” whether they received unemployment insurance in that wave). This question was only asked of individuals who did not report currently having a job (we adjusted for having a job in each survey wave in the analyses). The CARES Act $600 supplement to unemployment insurance should have been in place for the entire duration of the study, though it may have taken time for some states to deliver it to recipients. Initial unemployment insurance benefits payments were a lump sum retroactive to the start date of unemployment, so that the first payments would have been larger than subsequent payments.

### Outcomes

The first outcome of interest was food insecurity, coded as a binary variable based on the question, “In the past seven days, were you worried you would run out of food because of a lack of money or other resources?” We coded “yes” responses as 1, “no” responses as 0, and “unsure” responses as missing. The second outcome of interest was whether respondents reported eating less due to financial constraints based on the question “In the past seven days, did you eat less than you thought you should because of a lack of money or other resources?”

### Covariates

In all analyses, we included individual fixed effects to adjust for time-invariant individual characteristics. We included survey wave fixed effects to adjust for national secular trends in exposure to UI benefits and the outcomes of interest. We also adjusted for several time varying, self-reported covariates, including receiving a federal stimulus payment, receiving Supplemental Nutrition Assistance Program (SNAP) benefits in the month prior to survey, and employment status at the time of survey. The CARES Act included a one-time $1200 stimulus payment to each adult and $500 for each dependent child aged 16 years or younger for individuals earning less than $75,000, household heads earning less than $112,500, or married couples earning less than $150,000. Stimulus payments were coded as 1 after the first wave in which the respondent answered “yes” to the question, “In the past month, did you or anyone in your household receive any of the following government benefits? Economic stimulus funds.” \ We coded SNAP benefits as 1 or 0 for each wave based on the respondent’s answer of “yes” or “no” to the question, “In the past month, did you or anyone in your household receive any of the following government benefits? Supplemental Nutrition Assistance Program (SNAP or Food Stamps).” The inclusion of stimulus and SNAP benefits accounts for receipt of other services that may both be correlated with receipt of UI benefits and food insecurity. The inclusion of employment helps restrict model comparisons to unemployed persons who received UI versus unemployed persons who did not.

### Analysis

We first described the demographic characteristics and household composition of people in the sample and their reports of food insecurity and eating less.

We then used a difference-in-differences research design to compare changes in food insecurity and eating less before and after individuals received unemployment insurance relative to changes in the same outcomes over time among individuals with no change in unemployment insurance receipt during that period.^19^ The validity of difference-in-difference methods rests on the assumption that those receiving unemployment insurance in a given period would have had the same trends in food insecurity as those who did not receive unemployment insurance or received it in different periods.^19^ In addition a standard difference-in-differences model, we estimated a complementary “event study” model,^20,21^ in which we estimated the differences in the outcome associated with each time period before and after individuals reported first receiving unemployment insurance. In this specification, the primary exposure variables of interest were dummy variables for four-week periods relative to receiving the first unemployment insurance payment. Individuals in the sample who never received unemployment insurance were assigned zeros for all of the time period dummies.

The event study approach has two critical advantages for our research question. First, the flexible estimation strategy enables us to assess whether changes in food insecurity preceded the arrival of UI benefits. This could occur if individuals anticipated that they would receive UI benefits and smoothed consumption by borrowing. A change in food insecurity preceding UI receipt could also be evidence of reverse causality or residual confounding, e.g. if people experiencing food insecurity were more likely to apply for UI. The event study specification allows for a transparent assessment of the parallel pretrends assumption underlying difference-in-differences models, as it explicitly estimates trends in food insecurity outcomes across individuals receiving and not receiving UI prior to benefit receipt. Second, the event study specification allows the researcher to assess whether treatment effects vary over time, and helps reduce bias in situations where both this occurs and the timing of treatment (here, receipt of unemployment insurance benefits) varies.^20^ The event study approach also allowed us to evaluate whether the relationship between unemployment insurance and food insecurity differed with initial lump sum payments of retroactive unemployment insurance and later payments.

Our statistical model is presented in **Equation 1**, where *FI*_*it*_ is a binary indicator for food insecurity, *UI*_*it*_ is a binary indicator that switches from 0 to 1 if the individual begins receiving unemployment insurance, *S*_*it*_ is a binary indicator that switches from 0 to 1 if the individual receives the stimulus payment, *SNAP*_*it*_ is a binary indicator for receiving SNAP benefits in the past month, *I*_*i*_ is individual fixed effects, and *t*_*t*_ is period fixed effects. In the event study, we replaced *UI*_*it*_ with dummy variables indicating the number of periods prior to or following receipt of unemployment in insurance. We used linear regression due to evidence that logistic models can underestimate standard errors in the presence of fixed effects.^22^ We clustered standard errors by individuals to account for serial autocorrelation.^23^

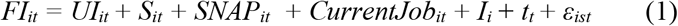

### Sensitivity Analyses

We conducted four sensitivity analyses. First, we ran the main difference-in-difference models restricted to individuals who participated in all six waves of the UCA survey. Second, we ran the main difference-in-difference models with survey weights designed to make the sample nationally representative. This analysis is informative as comparisons between weighted and unweighted models can help serve as a specification check.^24^ Third, we ran the main difference-in-differences models as logistic rather than linear models. Fourth, we ran the analysis in the subgroup of people living in households earning less than $20,000 per year to determine whether the association between unemployment insurance and food insecurity was concentrated among people in low-income households.

## Results

### Sample Characteristics

There were a total of 7120 participants in the UCA. Of 7002 participants who reported whether they were employed in February, 4503 (64%) were employed. Of those who were employed, 2361 (52.5%) had a household income of less than $75,000. Of these participants who met the sample inclusion criteria, 2185 (92.5%) participants responded to two or more UCA surveys and did not have missing data for covariates (Table 1, column 1). A total of 885 (40.5%) of those participants reported being unemployed during at least one wave of the UCA between April 1 and July 8, 2020 (Table 1, Column 2) and comprised the main sample. Participants responded to a mean of 5 of 6 UCA waves. The proportion of participants who reported not having a job in any given wave of the survey ranged from a high of 31.2% during the wave 2 (April 15 to May 12) survey to a low of 26.3% during the most recent, wave 6 (June 10 to July 8) survey. Participants who were Hispanic, non-white, lower income, and lesbian, gay, bisexual, or transgender (LGBT) were more likely to report unemployment.

**Table 1:** Demographic and household characteristics of participants in sample.

|  |  | Unemployment |  | Sample characteristics |  |
| --- | --- | --- | --- | --- | --- |
| Characteristic | (1)<br>N with<br>household<br>income<br><\$75,000 | (2)<br>N<br>unemployed<br>(Main<br>sample) | (3)<br>%<br>unemployed<br>at any point | (4)<br>% with<br>characteristic<br>in sample<br>(unweighted) | (5)<br>% with<br>characteristic<br>in sample<br>(weighted,<br>wave 5) |
| **Total** | 2185 | 885 | 40.5 | 100.0 | 100.0 |
| **Race and ethnicity** |  |  |  |  |  |
| Non-Hispanic White | 1259 | 473 | 37.6 | 53.5 | 52.6 |
| Non-Hispanic Black | 228 | 94 | 41.2 | 10.6 | 16.4 |
| Non-Hispanic American<br>Indian or Alaska Native | 26 | 9 | 34.6 | 1.0 | 0.5 |
| Non-Hispanic Asian | 121 | 53 | 43.8 | 6.0 | 5.5 |
| Non-Hispanic<br>Hawaiian/Pacific<br>Islander | 7 | 2 | 28.6 | 0.2 | 0.1 |
| Non- Hispanic Mixed<br>Race | 89 | 41 | 46.1 | 4.6 | 4.2 |
| Hispanic | 454 | 213 | 46.9 | 24.1 | 20.8 |
| **Sex** |  |  |  |  |  |
| Female | 1382 | 575 | 41.6 | 65.0 | 58.3 |
| Male | 803 | 310 | 38.6 | 35.0 | 41.7 |
| **Income group** |  |  |  |  |  |
| <\$20,000 | 350 | 214 | 59.8 | 24.2 | 25.6 |
| \$20,000 to \$29,999 | 295 | 145 | 56.2 | 16.4 | 17.2 |
| \$30,000 to \$39,999 | 376 | 155 | 43.5 | 17.5 | 16.9 |
| \$40,000 to \$59,999 | 680 | 238 | 38.6 | 26.9 | 26.5 |
| \$60,000 to \$74,999 | 484 | 133 | 30.6 | 15.0 | 13.7 |
| **Age group** |  |  |  |  |  |
| 18 to 29 years | 368 | 170 | 61.1 | 19.2 | 20.4 |
| 30 to 39 years | 522 | 191 | 49.2 | 21.6 | 26.5 |
| 40 to 49 years | 435 | 152 | 41.2 | 17.2 | 15.8 |
| 50 to 59 years | 456 | 183 | 40.1 | 20.7 | 18.6 |
| ≥60 years | 404 | 189 | 46.8 | 21.4 | 18.7 |
| **Sexual orientation** |  |  |  |  |  |
| Yes | 220 | 102 | 46.4 | 11.5 | 13.2 |
| No | 1965 | 783 | 39.8 | 88.5 | 86.8 |
| **Household composition** |  |  |  |  |  |
| Single adult, no children | 487 | 208 | 42.7 | 23.5 | 18.7 |
| Single adult, with<br>children | 90 | 38 | 42.2 | 4.3 | 5.6 |
| ≥2 adults, no children | 974 | 403 | 41.3 | 45.5 | 45.9 |
| ≥2 adults, with children | 634 | 236 | 37.2 | 26.7 | 29.9 |

Note: Characteristics are based on the first observation for each participant in the sample. All observations are from individuals who participated in at least 2 waves of the UCA survey. Numbers and percents represent the full sample and are unweighted in columns (1) – (4). Numbers in column 5 are weighted and reflect data from 778 participants in Wave 3 of the study, the wave in which the most participants in the sample participated.

There were a total of 885 people in the main sample of individuals living in households earning less than $75,000 in the past 12 months who had experienced unemployment between April 1 and July 8, 2020 and who participated in at least two waves of the UCA survey. Most (53%) were non-Hispanic White, 24% were Hispanic, 11% were non-Hispanic Black, and the remainder were of other races and ethnicities (Table 1, column 4). Those with higher household incomes made up less of the sample (15% made $60,000 to $74,999), whereas those earning less than $20,000 made up 24% of the sample. There were a similar proportion of people from each age group, and 12% reported being LGBT. More participants (46%) lived in households with two or more adults and no children, whereas 24% were adults who lived alone, 27% were two or more adults with children, and 4% were single adults with children.

A total of 341 (38.5%) of participants in the sample received unemployment insurance during the study period (Appendix Table 2). The greatest proportion of people who received unemployment insurance first received it in wave 2 (11.1%), while the least received it in wave 6 (2.8%).

### Food Insecurity

Overall, 31% of participants reported food insecurity and 33% reported eating less due to financial constraints during at least one wave of the survey (Table 2). During the April 1 to July 8 study period, the mean proportion of participants reporting food insecurity at any given time was 14.5% and the mean proportion of participants reporting eating less was also 14.5%. The greatest level of food insecurity was in the first wave of the survey (April 1 to April 28), when 21.9% of families reported food insecurity. Food insecurity declined with each wave of the survey, to 10.5% in the 6^th^ wave of the survey (June 10 to July 8). Food insecurity and eating less declined more among people who received unemployment insurance, but not to the level of those who remained employed (Appendix Figure 1). Food insecurity was trending up again among people who received unemployment insurance in the 6^th^ survey wave (June 10 to July 8). Participants in the sample who were Hispanic, non-white, younger, lower income, LGBT, and single adults living with children were more likely to report food insecurity and eating less.

**Table 2:** Ever reporting food insecurity or eating less by participant characteristics (N=885)

| Characteristic | Food insecurity (%) |  | Eating less (%) |
| --- | --- | --- | --- |
| **Total** | 31.1 |  | 32.7 |
| **Race and ethnicity** |  |  |  |
| Non-Hispanic White | 21.8 |  | 24.9 |
| Non-Hispanic Black | 34.0 |  | 38.3 |
| Non-Hispanic American Indian or Alaska Native | 33.3 |  | 22.2 |
| Non-Hispanic Asian | 33.9 |  | 43.4 |
| Non-Hispanic Hawaiian/Pacific Islander | Sample too small |  | Sample too small |
| Non- Hispanic Mixed Race | 39.0 |  | 39.0 |
| Hispanic | 48.4 |  | 44.1 |
| **Sex** |  |  |  |
| Female | 32.5 |  | 32.7 |
| Male | 28.4 |  | 32.6 |
| **Income group** |  |  |  |
| <\$20,000 | 49.1 | | 49.5 |
| \$20,000 to \$29,999 | 32.4 | | 31.7 |
| \$30,000 to \$39,999 | 31.0 | | 29.0 |
| \$40,000 to \$59,999 | 23.5 | | 29.4 |
| \$60,000 to \$74,999 | 14.3 | | 16.5 |
| **Age group** |  |  |  |
| 18 to 29 years | 41.8 |  | 44.1 |
| 30 to 39 years | 41.9 |  | 42.4 |
| 40 to 49 years | 38.8 |  | 42.1 |
| 50 to 59 years | 21.9 |  | 22.4 |
| ≥60 years | 13.2 |  | 14.8 |
| **LGBT** |  |  |  |
| Yes | 45.0 |  | 49.0 |
| No | 29.2 |  | 30.5 |
| **Household composition** |  |  |  |
| Single adult, no children | 30.8 |  | 31.7 |
| Single adult, with children | 60.5 |  | 57.9 |
| ≥2 adults, no children | 26.5 |  | 29.0 |
| ≥2 adults, with children | 35.2 |  | 35.6 |

Notes: Reflects the percent of participants who ever reported food insecurity or eating less due to a lack of money or resources by participant characteristic. Numbers and percents are unweighted.

**Figure 1.**
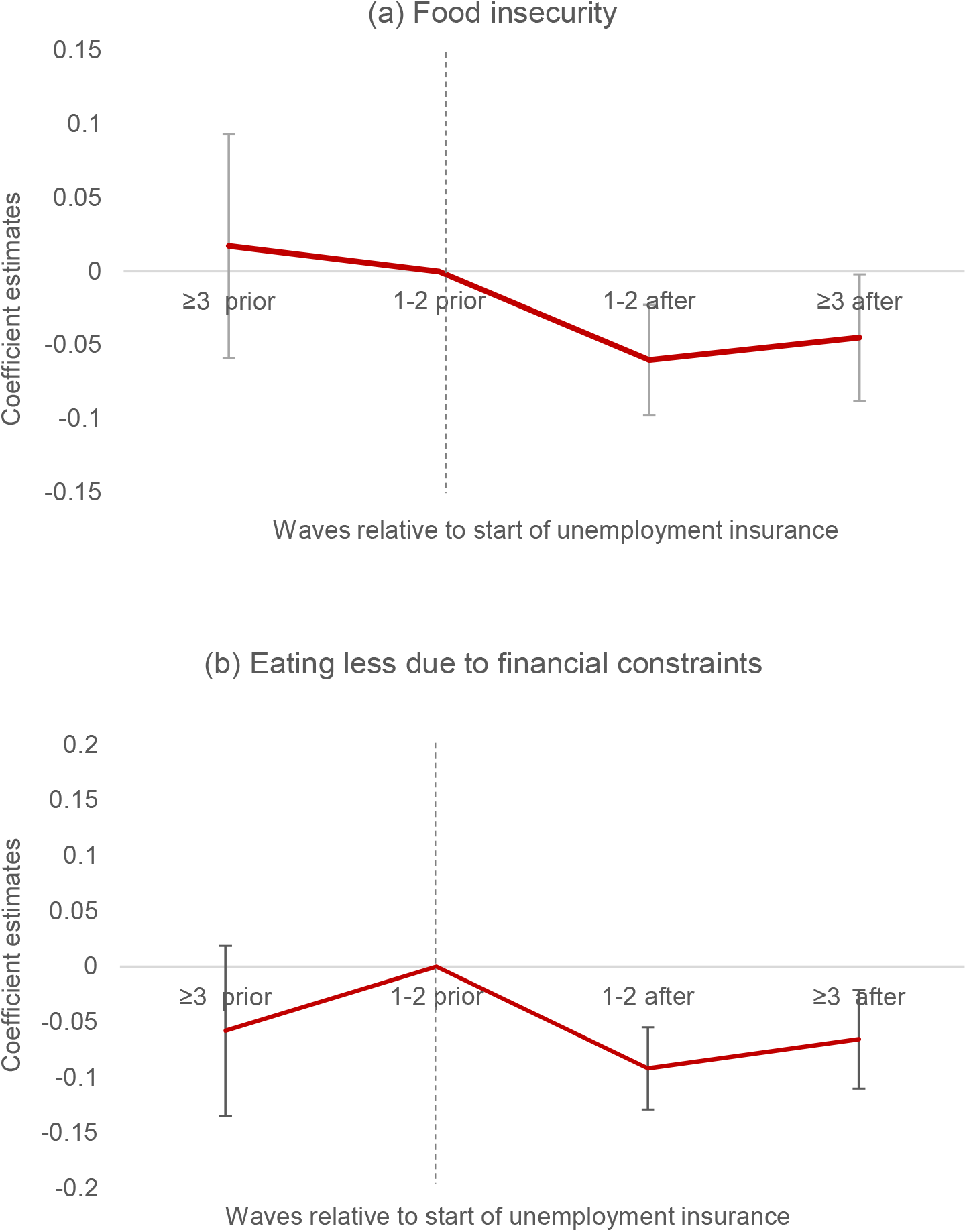
Event study estimates of the relationship between unemployment insurance and food insecurity and eating less. Notes: Event study estimates of food insecurity and eating less due to financial constraints. Coefficient estimates represent the estimated association between receiving unemployment insurance for the given number of periods and the food insecurity outcome. Estimated coefficients are based on running the main regression with all covariates and replacing the main unemployment insurance exposure variable with binary indicators for periods relative to receipt of unemployment insurance among those who received unemployment insurance, with individuals whose unemployment insurance status did not change during the study period set to zero. Estimates are adjusted for stimulus payments, receipt of SNAP benefits within the past 4 weeks, current employment status, and study wave, as well as individual-level fixed effects. The reference period is the period immediately prior to receipt of unemployment insurance. The dashed lined represents the period after which individuals received unemployment insurance. Standard errors are clustered by individual. The error bars represent 95% confidence intervals.

### Difference-in-differences estimates

We found that unemployment insurance was associated with a 4.4 percentage point (95% CI: −7.8 to −0.9 percentage points) decline in food insecurity overall (Table 3). The 4.4 percentage point overall decline in food insecurity is equivalent to a 30.3% relative decline from the average of 14.5% during the full study period. Unemployment insurance was also associated with a 6.1 percentage point (95% CI: −9.6 to −2.7 percentage point) decline in eating less. The 6.1 percentage point overall decline in eating less is equivalent to a 42% relative decline from the average of 14.5% during the full study period. Current employment was associated with a decline in eating less of 4.1 percentage points (95% CI: −6.7 to −1.4 percentage points) but was not associated with food insecurity (−1.7 percentage points, 95% CI: −4.3 to 0.9 percentage points). The federal stimulus payment (−1.42 percentage points, 95% CI: −4.7 to 1.9) and SNAP (−2.3, 95% CI: −8.9 to 4.4) were not associated with a change in food insecurity, nor were the federal stimulus payment (−1.4 percentage points, 95% CI: −5.1 to 2.3) or SNAP (4.0 percentage points, 95% CI: −2.6 to 10.6) associated with eating less.

**Table 3:**
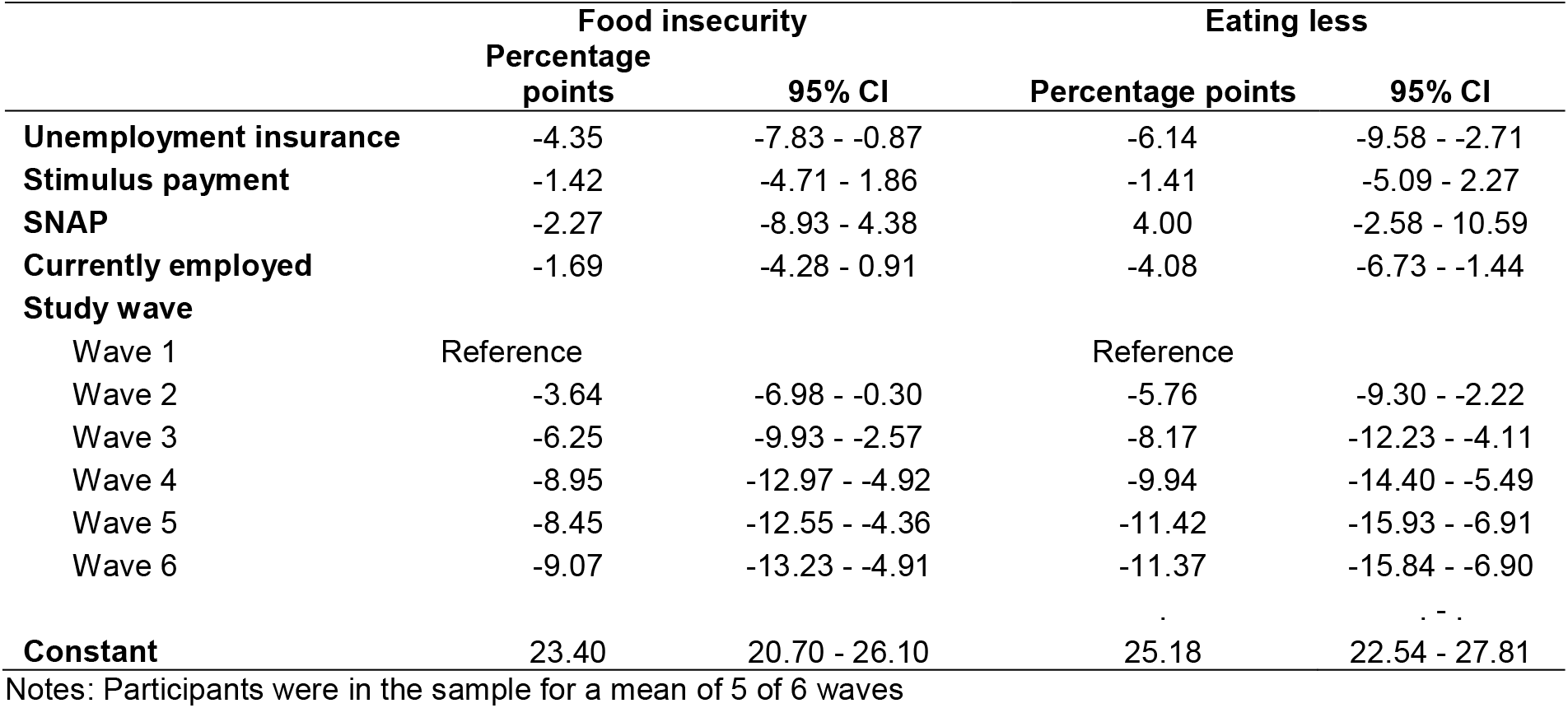
Main difference-in-differences estimates of the relationship between unemployment insurance and outcomes of food insecurity and eating less (N=885)

|  | Food insecurity |  | Eating less |  |
| --- | --- | --- | --- | --- |
|  | Percentage points | 95% CI | Percentage points | 95% CI |
| **Unemployment insurance** | -4.35 | -7.83 - -0.87 | -6.14 | -9.58 - -2.71 |
| **Stimulus payment** | -1.42 | -4.71 - 1.86 | -1.41 | -5.09 - 2.27 |
| **SNAP** | -2.27 | -8.93 - 4.38 | 4.00 | -2.58 - 10.59 |
| **Currently employed** | -1.69 | -4.28 - 0.91 | -4.08 | -6.73 - -1.44 |
| **Study wave** |  |  |  |  |
| Wave 1 | Reference |  | Reference |  |
| Wave 2 | -3.64 | -6.98 - -0.30 | -5.76 | -9.30 - -2.22 |
| Wave 3 | -6.25 | -9.93 - -2.57 | -8.17 | -12.23 - -4.11 |
| Wave 4 | -8.95 | -12.97 - -4.92 | -9.94 | -14.40 - -5.49 |
| Wave 5 | -8.45 | -12.55 - -4.36 | -11.42 | -15.93 - -6.91 |
| Wave 6 | -9.07 | -13.23 - -4.91 | -11.37 | -15.84 - -6.90 |
| **Constant** | 23.40 | 20.70 - 26.10 | 25.18 | 22.54 - 27.81 |

Notes: Participants were in the sample for a mean of 5 of 6 waves

Results from event study analyses were similar. The results (Figure 1) demonstrated that reductions in food insecurity and eating less were greatest in the four-week period immediately following receipt of unemployment insurance. There was a 6.0 percentage point (95% CI: −9.8 to −2.3 percentage points) decline in food insecurity in the four-week period immediately following first receipt of unemployment insurance (event study in Figure 1a), equivalent to a 41% relative decline. There was a 9.2 percentage point (95% CI: −12.9 to −5.5 percentage points) decline in eating less in the four-week period immediately following the first receipt of unemployment insurance (event study in Figure 1b), equivalent to a 63% relative decline.

The event study estimates do not suggest the presence of differential trends in food insecurity or eating less prior to receipt of UI benefits. Declines in food insecurity and eating less prior to receipt of unemployment insurance were similar for those who received unemployment insurance in different waves (Appendix Figure A2).

### Sensitivity analyses

Sensitivity analyses were consistent with the main result for individuals who participated in all six waves of the UCA survey (Appendix Table 3), models with survey weights (Appendix Table 4) and for logistic rather than linear models (Appendix Table 5). As anticipated, effect size estimates were larger for those living in households with income of less than $20,000 than for the full sample with income of less than $75,000 (Appendix Table 6).

## Discussion

In this national study, we found that receipt of unemployment insurance was associated with a 30% reduction in food insecurity and a 42% decline in eating less due to financial constraints among people with household earnings of less than $75,000 who lost their jobs during the COVID-19 pandemic. Results were larger for those in the lowest income group. There was a $600 weekly federal supplement to unemployment insurance in place during the full study period that may have contributed to reductions in food insecurity. Our findings that unemployment insurance was associated with reduced food insecurity, particularly in the period immediately following first receipt of unemployment insurance, is consistent with other work showing that household spending increased immediately following receipt of unemployment insurance for households that began receiving unemployment insurance in April 2020.^25^ Our findings are also consistent with prior evidence that unemployment insurance is associated with reduced food insecurity.^15^ Unemployment insurance did not reduce food insecurity to the level of those who remained employed, and other research has found that the $600 weekly federal supplement to unemployment insurance did not reduce employment among people receiving unemployment insurance; in the current US context in which many businesses are operating at much lower capacity than prior to the COVID-19 pandemic, the federal supplement to unemployment insurance may contribute to reduced food insecurity without reducing employment.

We found that 41% of people living in households with incomes below $75,000 who were employed in February 2020 reported being unemployed at some point between April 1 and July 8 of 2020. Among those in households with incomes below $75,000 who were unemployed at any point, 31% reported food insecurity and 33% reported eating less due to financial constraints. Food insecurity was particularly pronounced among those with lower incomes, who were Hispanic, who were LGBT, and who were single parents. Food insecurity declined over time for both those who did and did not receive unemployment insurance, but declined more for those who received unemployment insurance. Notably, food insecurity started to increase again among those who received unemployment insurance in the final wave of the survey, conducted June 10 to July 8, with the increase in food insecurity potentially due to the $600 federal supplement to unemployment insurance ending at the end of July. Our estimates of food insecurity are consistent with estimates from the Census Pulse survey, which indicated that 23% of US households across all income levels experienced food insecurity between April 23–May 19.^12^

While the federal government provided a supplement to unemployment insurance, it is states that manage unemployment insurance programs. Eligibility for unemployment insurance, the base period used to calculate benefits amounts, and requirements for those receiving unemployment insurance continue to vary widely from state to state,^14^ affecting the proportion of people receiving unemployment insurance and the amount that people are receiving. In March 2020, the proportion of unemployed people who were receiving unemployment insurance ranged from 7.6% in Florida to 65.9% in Massachusetts.^26^ The amount of unemployment insurance without the federal supplement that expired in July ranged from $240 per week in Arizona to $820 per week in Massachusetts.^14^ State, as well as federal, policies will continue to play an important role in shaping unemployment insurance and associated food insecurity, health, and well-being.

### Limitations

As with all difference-in-differences analyses, particularly those conducted in the rapidly changing policy context of COVID-19,^27^ our study has clear limitations. Unemployment insurance, stimulus payments, and SNAP were often delivered in close temporal proximity to each other, making it difficult to fully distinguish the effects of each, even after covariate adjustment. We were also unable to distinguish the effect of the additional $600 in federal unemployment benefits from the benefits of receiving any unemployment insurance because the supplement was in place for the entire duration of the study. In addition to these statistical concerns, measures of both the outcome and exposure rely on self-report, which may be prone to bias.

We investigated why the estimates of the proportion of people who were employed in February who lost their jobs are greater than the 11.1% unemployment estimates reflected in the Bureau of Labor Statistics Employment Situation Summary Report.^28^ The BLS estimates may be underestimates due to misclassification bias,^29^ and our estimates are consistent with the Federal Reserve Bank’s estimates that 20% of all those who were employed in February 2020 and 39% of those with household incomes of less than $40,000 were unemployed in March 2020.^30^

## Conclusion

More than 40% of people with household incomes less than $75,000 who were employed in February 2020 lost their jobs during the COVID-19 pandemic. Among those who lost their jobs, 31% reported food insecurity and 33% reported eating less due to financial constraints between April 1 and July 8, 2020. During the period when the CARES Act provided a $600 supplement to state unemployment insurance, receiving unemployment insurance was associated with a 30% reduction in reporting any food insecurity and a 42% decline in eating less. Policymakers may wish to consider continued investment in unemployment insurance as an approach to reducing food insecurity in the context of high unemployment during the continuing COVID-19 pandemic.

## Data Availability

The data are publicly available from the University of Southern California

https://uasdata.usc.edu/index.php

## Acknowledgments

The study was funded by the Boston University Clinical and Translational Science Institute, National Institutes of Health grant UL1TR001430.

## Notes

### Competing Interest Statement

The authors have declared no competing interest.

### Funding Statement

The study was funded by the Boston University Clinical and Translational Science Institute grant UL1TR001430

### Author Declarations

The analysis was based on analyses of deidentified secondary data and is considered not human subjects research

### Summary of Updates

Revised abstract to make more concise and improved clarity of appendix figures

